# Co-designed and co-delivered place-based community interventions to reduce inequity in early initiation of antenatal care: Findings from the cluster randomised controlled Community REACH trial

**DOI:** 10.1101/2024.11.15.24317381

**Authors:** A Harden, M Wiggns, L Sweeny, C Salisbury, T Hamborg, S Eldridge, L Greenberg, R Hunter, E Bordea, McCourt, B Hatherall, G Findlay, A Renton, R Ajayi, C Durham, A Adeyemo, B Harvey, K Mondeh, V VanLessen

## Abstract

**Background:** Early initiation of antenatal care provides timely screening, advice and support. Inequities in early care initiation exist in high income countries, but there is scant evidence on effective interventions. The Community REACH trial aimed to assess the effectiveness of co-produced place-based interventions to strengthen community support for early care initiation.

**Methods:** Matched-pair cluster randomised trial in socially disadvantaged and ethnically diverse areas in England. Electoral wards with low rates of early care initiation were matched and randomly allocated to intervention or control (usual care) (n=10 pairs). Following a three month co-design phase, community organisations and volunteers in intervention sites conducted targeted outreach activities over six months. The primary outcome was initiation of antenatal care by the 12^th^ completed week of pregnancy.

**Results:** There was no evidence of a difference in the primary outcome (OR 1.07(0.89; 1.28)). There were also no statistically significant differences in rates of emergency caesarean, pre-term birth, low birth weight, smoking, or breastfeeding. There was a higher rate of care initiation by 10 weeks and fewer antenatal admissions in the intervention arm during the intervention period although differences were not sustained after it finished.

**Conclusion:** This rigorous evaluation found limited impact of short-term place-based interventions to strengthen community support for early initiation of antenatal care. Future initiatives may benefit from embedding in integrated health and care structures to ensure sufficient time and resources for mobilisation of community assets and focusing on smaller ‘hyper-local’ neighbourhoods. Actions to tackle wider structural and organisational barriers are also needed.

**Trial registration:** ISRCTN registry: registration number 63066975. Registered on 18 August 2015.

WHAT IS ALREADY KNOWN ON THIS TOPIC

- Previous research in high income countries has identified inequalities in access to antenatal care, yet there is little evidence on interventions to improve early initiation of antenatal care.
- Co-produced place-based interventions which develop and strengthen community support offer a promising approach to tackle health inequalities but this type of approach has not yet been rigorously tested in the context of early initiation of antenatal care in high income countries.

WHAT THIS STUDY ADDS

- It was possible to develop and implement a co-designed and co-delivered place-based six month intervention to increase early access to antenatal care in ethnically and linguistically diverse inner-city neighbourhoods.
- There is no evidence that this short-term intervention increased the proportion of women accessing antenatal care before 12 completed weeks of pregnancy; initiation of care by 10 weeks increased significantly whilst the intervention was running, but this was not maintained after it ended.
- Intervention implementation varied across sites in the extent to which it reflected the underpinning intervention logic of co-production and community development; there was, however, no evidence of an association between implementation and intervention effect estimates.

HOW THIS STUDY MIGHT AFFECT RESEARCH, PRACTICE OR POLICY

- Given the results of this study, funders and intervention developers of future initiatives to strengthen community support should ensure a longer lead in time so that interventions can be embedded within integrated health and care structures and allow sufficient time, resources and capability for co-production, the development of community relationships and asset-based ways of working.
- Efforts to tackle inequalities in early initiation of antenatal care should also consider service-level actions such as geographical location, availability of interpreters and anti-racist practice training.

## Introduction

Reducing inequities in maternal and infant health outcomes is a priority for public health policy worldwide. In high income countries such as the UK, socially disadvantaged groups and those from ethnically minoritised backgrounds continue to experience the worst outcomes [1]. Antenatal care, the package of care provided from conception to the onset of labour including components such as health promotion and screening, has been shown to prevent adverse outcomes including maternal and neonatal mortality [2-4]. Starting antenatal care within early pregnancy is recommended to receive the greatest benefit [2,5]. There are, however, inequities in care initiation with later access linked to socio-economic deprivation and more likely amongst minoritised ethnic groups [6-11].

Systematic reviews have found a dearth of evidence on what works to increase early initiation of antenatal care [10, 12, 13]. Evidence on barriers and facilitators amongst those more likely to experience late access highlight the complexity of navigating health systems and a lack of promotion of the importance of early initiation of care [12, 8, 14]. In high income countries with universal free access to health services, these reviews suggest as promising approaches in which maternity services collaborate with community organisations to promote early initiation in proactive, accessible and culturally safe ways. In low and middle-income countries participatory strategies involving peer leaders, advocacy and community health committees have been shown to promote earlier initiation of care [15,16].

Place-based interventions which target the physical, economic or socio-cultural aspects of place offer a promising approach for reducing health inequalities [17, 18]. Those that aim to change socio-cultural aspects of a place (e.g. community cohesion, support networks) fall into the ‘strengthening communities’ category of Whitehead’s typology of actions to reduce inequalities in health [19]. It is imperative that marginalised and underserved communities are enabled to co-produce interventions that target them [20] and there is emerging evidence that when this happens interventions are more likely to be effective [21,22]. The effectiveness of co-produced place-based interventions which strengthen community support has not yet been rigorously tested regarding early initiation of antenatal care in high income countries. This paper describes a study to address this gap.

The aims of this study’s intervention were to a) raise awareness in local communities of the value of antenatal care and its early uptake and how to access it, and b) activate community assets and the wider health and care system to support early access. The longer-term aim was to change local social norms to sustain any increase in early initiation of antenatal care.

## Methods

### Trial Design

Two-arm, matched-pair repeated cross-sectional trial, conducted in ethnically and linguistically diverse inner-city electoral wards^1^ in South East England [23]. The study included process and economic evaluations the results of which are presented elsewhere [24, 25]. For the trial protocol see [23].

### Intervention

The intervention logic model was informed primarily by the concepts of co-production, community development and health literacy (Figure 1). It was also informed by developmental work to examine barriers to early access amongst ethnically diverse and socially disadvantaged communities [6, 8] and the Well Communities framework for improving health in disadvantaged neighbourhoods [26]. Well Communities specialists and a social design agency ran co-design workshops with residents and health and care system staff in each intervention site over a three-month period. Intervention ideas were pooled and formed an intervention plan with several components.

**Figure 1:**
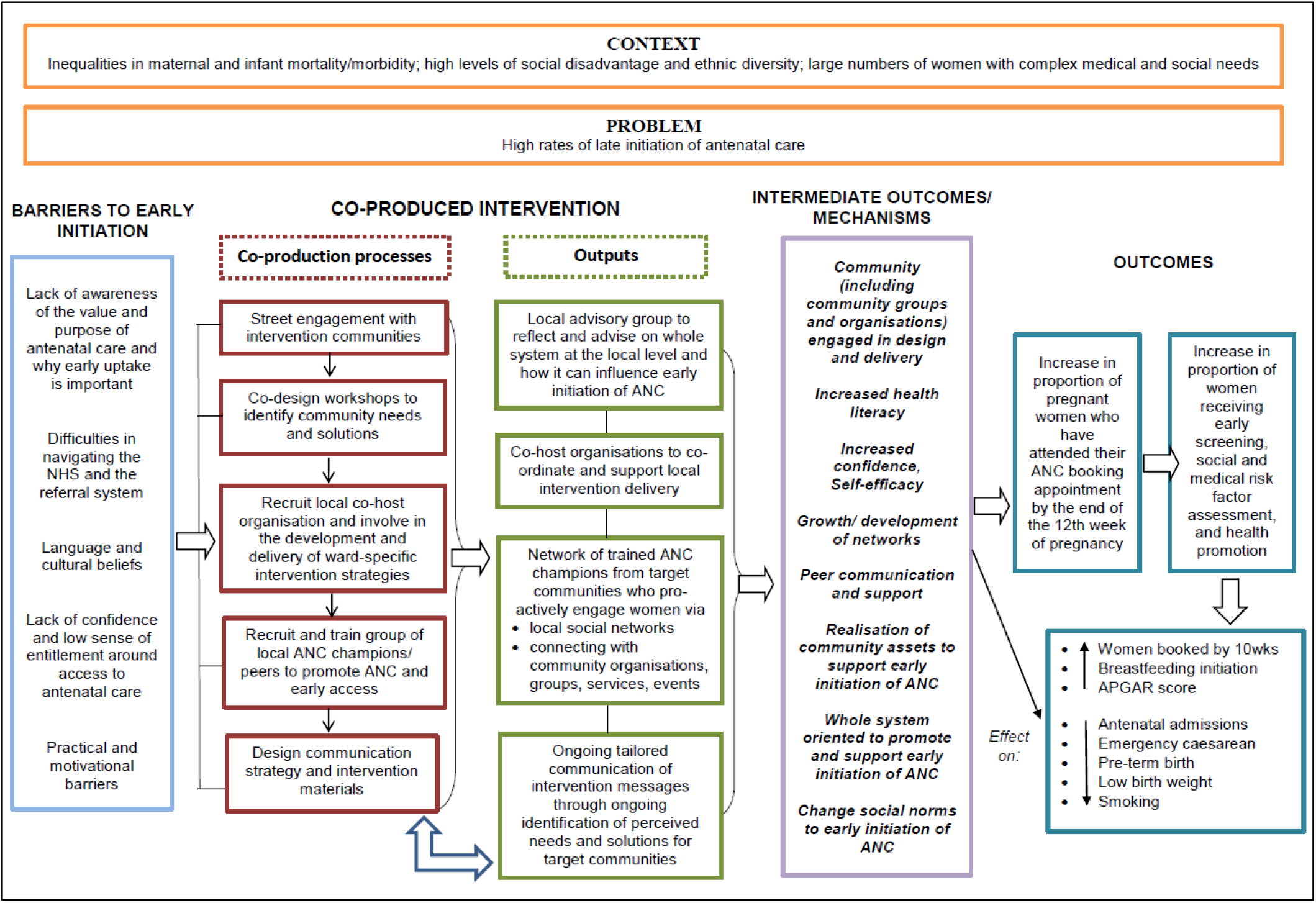
The Community REACH intervention logic model. (*reproduced from Sawtell et al. 2018*)

In each intervention area, a local community organisation managed the intervention over nine months (three months set up; six months intervention delivery). They convened local advisory groups to advise on key issues restricting early initiation of antenatal care and possible solutions. They recruited and trained community members as ‘antenatal champions’ to conduct conversations with people in their communities about the benefits of early care initiation and how to access care. The intervention co-ordinator and antenatal champions also held targeted conversations with community leaders, local health and care services (e.g. GP practices) and community assets (e.g. libraries, places of worship, local businesses such as barbershops) to generate community support for early care initiation. Further details are provided in Supplementary File 1 and in [27]).

### Recruitment and randomisation

Six NHS Trusts were recruited between April-November 2015. The unit of randomisation was electoral wards served by these Trusts. Wards with high delayed rates of initiation of antenatal care (<90% rate of antenatal booking by the end of the 12th completed week of pregnancy) were considered for inclusion into the study. The number of potential study wards was further reduced by removing wards so that no wards neighboured one another to reduce likelihood of contamination. Ward pairs were matched on antenatal care initiation rates (low or very low), using data from a 6-month pre-trial period. Within each pair one ward was randomised to intervention and the other one to control (usual care) implying a 1:1 allocation ratio. Randomisation was conducted remotely by the Pragmatic Clinical Trials Unit at Queen Mary, University of London.

### Outcomes

The primary outcome was antenatal booking appointment attendance by the end of the 12th completed week of pregnancy (12 weeks and 6 days) as a binary (yes/no) variable. Secondary outcomes included: Attendance at antenatal booking appointment by 10 weeks +0 days of pregnancy; Number of antenatal hospital admissions; Smoking during pregnancy; Proportion of Emergency Caesarean deliveries; Proportion of pre-term births; Weight of baby at delivery; Smoking at time of birth; Infant feeding method at discharge from hospital.

### Data collection

All outcomes were collected via routine maternity data, provided electronically by data informatics teams from NHS Trusts participating in the study. In this repeated cross-sectional study design, outcome data were obtained from three different cohorts of women: cohort 1 (baseline) was prior to any intervention activities; cohort 2(FU1) was used to assess the treatment effect from 1 month after intervention commencement and cohort 3 (FU2) was used to assess the sustained effect of the intervention after implementation ended. The period length for all 3 cohorts was 6 months. Time periods determining cohorts for control sites were the same as for their paired intervention site.

Follow-up data were extracted for the period June 2017 to October 2019.

### Sample size

The trial was designed to detect an increase in antenatal booking by 12 weeks and 6 days gestation from 73% to 80%, with 90% power at the 5% significance level, accounting for clustering by ward (ICC = 0.005) and assuming a mean cluster size of 130 and matching correlation = 0.3. According to this sample size calculation nine clusters in both intervention and control groups are required, which equated to at least 798 women per trial arm. To guard against loss of power if a cluster was lost, one cluster was added to each group leading to a total recruitment target of 20 clusters.

### Statistical analysis

#### a) Main analysis

The primary outcome was analysed using a two-stage individual participant data meta-analysis technique for analysing paired cluster randomised controlled trials. Each matched pair of sites was regarded as an individual study in a meta-analysis for which the odds ratio was estimated. These odds ratios were subsequently combined using a random effects model. Restricted maximum likelihood estimation was used. The Hartung-Knapp modification to the DeSimonian-Laird estimator of the between study variance was used to construct t-based 95% confidence intervals for effect estimators. All secondary outcomes were analysed using the same principles and analysis approach as the primary outcome except for the outcome ‘Number of antenatal admissions’. The use of the primary outcome model for count variables has not been described in the literature. This outcome was therefore analysed using a paired t-test on the mean number of admissions per site.

The first appearance/appointment of a women in hospital within the routine maternity hospital data, up until and including birth, is used as the primary outcome value. The assumption was therefore that the amount of missing data would be negligible (<2%) and a blinded interim assessment of missingness confirmed this. The main analysis is therefore conducted on the available data without imputation.

#### b) Sub-group analysis

Subgroup analyses were planned on pre-defined subgroups (first vs subsequent pregnancy; ethnicity, deprivation, baseline booking rate and intervention model) for hypothesis generation, recognising that the study has limited power to detect differences at sub-group level. All subgroup analyses were performed by estimating the treatment-covariate interaction within each pair of sites and pooling of the effect estimates thereafter.

Process data were used to assess intervention implementation [24]. Each intervention site was assessed for: 1) how closely implementation aligned with the community development ethos reflected in the logic model (scored high/ med/ low); and 2) the number of reported conversations between antenatal champions and targeted community members (high/med/low). This led to the construction of two models of implementation which were used in subgroup analysis:

- ***Model A***: sites where implementation had more focus on numbers of conversations with community members and less focus on wider/more embedded community development.
- ***Model B***: sites where implementation was more concentrated on embedded community development, but fewer conversations were reported.

#### c) Sensitivity analyses

The following analyses were carried out:

1. The treatment effect was estimated leaving out any site in which the intervention was not fully delivered as intended (per protocol analysis)
2. The treatment effect was estimated for the primary outcome using a model adjusted for individual-level covariates IMD and ethnicity as fixed effects. Adjusted marginal proportion for treatment groups were estimated using the approach by Norton et al [28].
3. Imputation of missing primary outcome values under different missing value assumptions.

#### d) Safety analysis

Maternal and infant deaths were analysed as safety measures across trial arms at each timepoint.

The full statistical analysis plan, with dates of recruitment and follow up, is publicly available at https://osf.io/cgfjw/.

### Ethics and consent to participate

This study conformed to the Declaration of Helsinki principles and was approved by the NHS Health Research Authority National Research Ethics Committee North East-York (27 March 2015, ref.15/NE/0106). All information collected during the trial was kept confidential and adhered to the 1998 Data Protection Act. There was no individual consent process for the collection of outcome data due to the reliance on anonymous routine data. Informed consent was sought from participants in the qualitative process evaluation which is reported elsewhere [24].

### Patient and public involvement

Members of the public were involved in the trial in multiple ways. The trial was part of a programme grant which included two lay co-investigators on the investigator group; they are also co-authors on this paper. Residents from intervention wards were involved in the co-design and delivery of the intervention and local community organisations managed intervention delivery.

## Results

### Study flow and participation

The consort diagram (Figure 2) indicates that all ten sites allocated to the intervention arm participated, with programmes delivered in nine sites for the full intervention period of six months; in one site, delivery was for three months due to intervention set-up delays.

**Figure 2:**
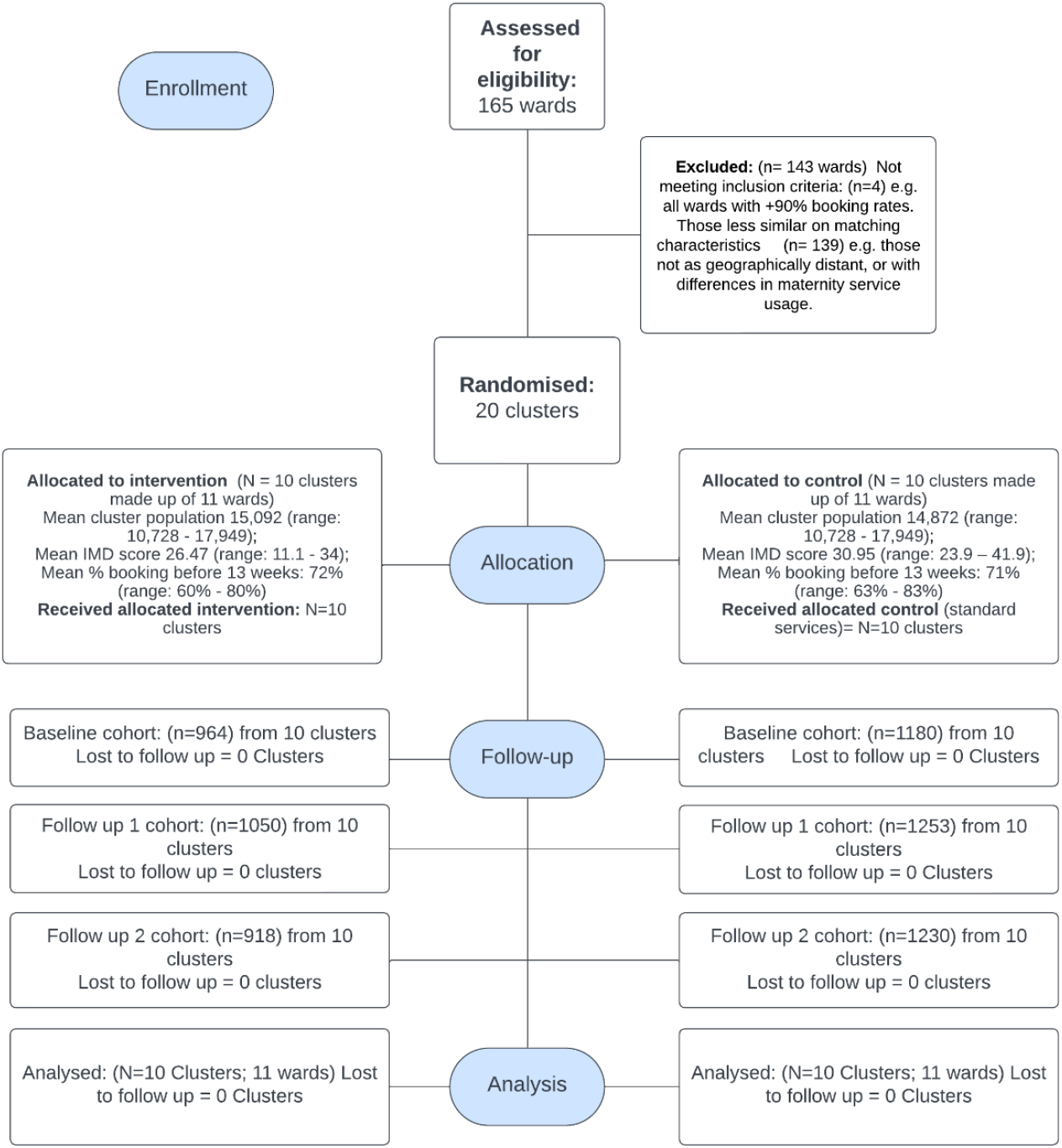
Community REACH trial consort diagram.

Data at the three time periods were provided for all intervention and control sites, and analysis was conducted with data from all sites.

### Balance between intervention and control sites

There was relatively even balance between wards allocated to the two arms in terms of population size and booking rates before 13 weeks of pregnancy at baseline (Figure 2). The wards within control sites were more deprived, with higher mean IMD scores than those within intervention sites (Figure 2). Demographic data for the three cohorts also flagged that more of those in control sites lived in areas of greater deprivation. Other demographics were evenly balanced with, for example, a high proportion of women from minoritised ethnic groups in both arms (Table 1).

**Table 1:** Baseline characteristics.

|  | Baseline cohort |  | First follow up cohort |  | Second follow-up cohort |  |
| --- | --- | --- | --- | --- | --- | --- |
|  | Intervention<br>(n=964) | Control<br>(n=1180) | Intervention<br>(n=1050) | Control<br>(n= 1253) | Intervention<br>(n=918) | Control<br>(n=1230) |
| Women's age at booking N(%) |  |  |  |  |  |  |
| <20 | 10 (1.34%) | 33 (3.36%) | 29 (3.32%) | 31 (2.80%) | 18 (1.96%) | 42 (3.41%) |
| 20-35 | 573 (76.71%) | 770 (78.49%) | 667 (76.32%) | 843 (76.01%) | 700 (76.25%) | 930 (75.61%) |
| >35 | 164 (21.95%) | 178 (18.14%) | 178 (20.37%) | 235 (21.19%) | 200 (21.79%) | 258 (20.98%) |
| missing | 217 (22.51%) | 199 (16.86%) | 176 (16.76%) | 144 (11.49%) | 0 (0.00%) | 0 (0.00%) |
| Parity at booking N(%) |  |  |  |  |  |  |
| 0 | 312 (36.19%) | 404 (37.86%) | 264 (30.70%) | 344 (33.14%) | 218 (28.24%) | 316 (29.51%) |
| 1-2 | 425 (49.30%) | 486 (45.55%) | 472 (54.88%) | 497 (47.88%) | 453 (58.68%) | 550 (51.35%) |
| 3+ | 125 (14.50%) | 177 (16.59%) | 124 (14.42%) | 197 (18.98%) | 101 (13.08%) | 205 (19.14%) |
| missing | 102 (10.58%) | 113 (9.58%) | 190 (18.10%) | 215 (17.16%) | 146 (15.90%) | 159 (12.93%) |
| Ethnicity N(%) |  |  |  |  |  |  |
| White British | 180 (17.34%) | 171 (12.89%) | 177 (16.43%) | 188 (13.83%) | 182 (18.24%) | 184 (14.06%) |
| White (other) | 209 (20.13%) | 355 (26.75%) | 219 (20.33%) | 365 (26.86%) | 250 (25.05%) | 395 (30.18%) |
| Asian | 134 (12.91%) | 179 (13.49%) | 175 (16.25%) | 198 (14.57%) | 218 (21.84%) | 223 (17.04%) |
| Black | 103 (9.92%) | 177 (13.34%) | 107 (9.94%) | 190 (13.98%) | 111 (11.12) | 215 (16.42%) |
| Other | 101 (9.73%) | 117 (8.82%) | 102 (9.47%) | 115 (8.46%) | 94 (9.42%) | 118 (9.01%) |
| Declined/Unknown | 94 (9.06%) | 129 (9.72%) | 120 (11.14%) | 159 (11.70%) | 143 (14.33%) | 174 (13.29%) |
| missing | 217 (22.51%) | 199 (16.86%) | 177 (16.86%) | 144 (11.49%) | 0 (0.00%) | 0 (0.00%) |
| Speaks English N(%) |  |  |  |  |  |  |
| Yes | 199 (51.96%) | 266 (55.53%) | 225 (46.49%) | 304 (49.92%) | 227 (39.48%) | 325 (43.45%) |
| missing | 581 (60.27%) | 701 (59.41%) | 566 (53.90%) | 644 (51.40%) | 343 (37.36%) | 482 (39.19%) |
| Deprivation quintile N(%) |  |  |  |  |  |  |
| 1-2 | 371 (38.49%) | 644 (54.58%) | 402 (38.29%) | 680 (54.27%) | 373 (40.63%) | 689 (56.02%) |
| 3-4 | 430 (44.61%) | 481 (40.76%) | 487 (46.38%) | 514 (41.02%) | 415 (45.21%) | 490 (39.84%) |
| 5-6 | 93 (9.65%) | 51 (4.32%) | 93 (8.86%) | 54 (4.31%) | 80 (8.71%) | 46 (3.74%) |
| 7-8 | 40 (4.15%) | 4 (0.34%) | 45 (4.29%) | 5 (0.40%) | 28 (3.05%) | 5 (0.41%) |
| 9-10 | 30 (3.11%) | 0 (0%) | 23 (2.19%) | 0 (0%) | 22 (2.40%) | 0 (0%) |
| missing | 0 (0.00%) | 0 (0.00%) | 0 (0.00%) | 0 (0.00%) | 0 (0.00%) | 0 (0.00%) |
| ToC after 12wk+6d N(%) |  |  |  |  |  |  |
| Yes | 45 (4.46%) | 117 (9.02%) | 42 (3.85%) | 106 (7.80%) | 57 (5.85%) | 85 (6.46%) |
| missing | 440 (43.61%) | 426 (32.85%) | 345 (31.59%) | 343 (25.24%) | 213 (21.85%) | 210 (15.97%) |
| Smoking at booking N(%) |  |  |  |  |  |  |
| Yes | 53 (7.24%) | 69 (7.09%) | 69 (8.23%) | 85 (8.00%) | 55 (6.46%) | 100 (8.69%) |
| missing | 232 (24.07%) | 207 (17.54%) | 212 (20.19%) | 190 (15.16%) | 67 (7.30%) | 79 (6.42%) |
| Risk category N(%) |  |  |  |  |  |  |
| Standard | 396 (53.88%) | 495 (50.98%) | 408 (40.22%) | 549 (50.27%) | 454 (50.56%) | 577 (47.69%) |
| Intermediate | 268 (36.46%) | 364 (37.49%) | 341 (39.47%) | 437 (40.02%) | 356 (39.64%) | 500 (41.32%) |
| Intensive | 71 (9.66%) | 112 (11.53%) | 115 (13.31%) | 106 (9.71%) | 88 (9.80%) | 133 (10.99%) |
| missing | 229 (23.76%) | 209 (17.71%) | 186 (17.71%) | 161 (12.85%) | 20 (2.18%) | 20 (1.63%) |

### Outcomes findings

#### a) Main results for primary and secondary outcomes for follow-up 1 cohort

There was no statistically significant difference between arms in the primary outcome (initiation of antenatal care by the end of the 12th completed week of pregnancy) in the follow-up 1 cohort in which outcomes were measured during the active implementation of the six-month intervention (I=83.06%, C=82.46%; OR1.07(0.89; 1.28) p=0.440) (Table 2). There were also no significant differences by trial arm for five of the seven secondary outcomes. There were similar rates of emergency caesarean, pre-term birth, low birth weight, smoking at discharge, and breastfeeding initiation in both the intervention and control arm (Table 2). There was, however, a higher rate of initiation of antenatal care by 10 weeks in the intervention arm (I= 49.47%; C= 40.14%; OR 1.68 (1.03; 2.75) p=0.041). The mean number of antenatal hospital admissions was also lower in the intervention arm (I= 0.47, C= OR -0.13 (−0.24; 0.01) p=0.030). (Table 2). Sensitivity analyses suggest that the intervention effect estimate of the primary outcome is robust (Supplementary file 2).

**Table 2:** Main results for primary and secondary outcomes.

| Follow up 1 Cohort |  |  |  |  |  |  |  |
| --- | --- | --- | --- | --- | --- | --- | --- |
|  | Included in analysis |  | Summary measure |  | Treatment effect |  |  |
|  | Intervention N (% miss) | Control N (% miss) | Intervention N (%) | Control N (%) | OR (95% CI) | OR p-value | RD (95% CI) |
| **Primary outcome** |  |  |  |  |  |  |  |
| Proportion of women having first antenatal appointment within 12wk + 6d | 1039 (1.05%) | 1243 (0.80%) | 863 (83.06%) | 1025 (82.46%) | 1.07 (0.89; 1.28) | 0.440 | 0.01 (-0.01; 0.04) |
| **Secondary outcomes** |  |  |  |  |  |  |  |
| Proportion of participants having first antenatal appointment within 10 wk+0d | 1039 (1.05%) | 1243 (0.80%) | 514 (49.47%) | 499 (40.14%) | 1.68 (1.03; 2.75) | 0.041 | 0.10 (0.01; 0.19) |
| No of hospital admissions (mean, med, (sd)) | 1012 (3.62%) | 1212 (3.27%) | 0.47, 0, (0.82) | 0.56, 0 (1.00) | -0.13 (-0.24; 0.01) | 0.030 | NA |
| Pre-term birth (<37 weeks) ('no') | 859 (17.56%) | 1003 (18.65%) | 783 (91.15%) | 917 (91.43%) | 0.95 (0.73; 1.24) | 0.692 | -0.00 (-0.02; 0.02) |
| Emergency Caesareans section ('no') | 876 (15.93%) | 1018 (17.44%) | 714 (81.51%) | 828 (81.34%) | 0.99 (0.65; 1.52) | 0.966 | 0.00 (-0.06; 0.06) |
| Low birth weight ('no') | 648 (37.81%) | 714 (42.09%) | 603 (93.06%) | 657 (92.02%) | 1.11 (0.76; 1.63) | 0.523 | 0.01 (-0.01; 0.04) |
| Smoking at discharge ('no') | 755 (28.10%) | 899 (28.25%) | 701 (92.85%) | 831 (92.44%) | 1.23 (0.74; 2.05) | 0.383 | 0.01 (-0.02; 0.05) |
| Initiated breastfeeding ('yes') | 786 (25.14%) | 934 (25.46%) | 711 (90.46%) | 835 (89.40%) | 1.28 (0.79; 2.08) | 0.276 | 0.02 (-0.02; 0.06) |

**Table 2: (cont'd) Main results for primary and secondary outcomes**
| Follow up 2 cohort |  |  |  |  |  |  |  |
| --- | --- | --- | --- | --- | --- | --- | --- |
|  | Included in analysis |  | Summary measure |  | Treatment effect |  |  |
|  | Intervention<br>N (% miss) | Control N (%<br>miss) | Intervention N (%) | Control N (%) | OR (95% CI) | OR<br>p-value | RD (95% CI) |
| **Primary outcome** |  |  |  |  |  |  |  |
| Proportion of women having first antenatal appointment within 12wk + 6d | 904 (1.53%) | 1209 (1.71%) | 708 (78.32%) | 955 (78.99%) | 0.98 (0.74; 1.30) | 0.888 | -0.00 (-0.05; 0.04) |
| **Secondary outcomes** |  |  |  |  |  |  |  |
| Proportion of participants having first antenatal appointment within 10 wk+0d | 904 (1.53%) | 1209 (1.71%) | 408 (45.13%) | 493 (40.78%) | 1.23 (0.85; 1.78) | 0.230 | 0.04 (-0.03; 0.11) |
| No of hospital admissions (mean, med, (sd)) | 885 (2.66%) | 1185 (2.94%) | 0.50,0, (1.00) | 0.55,0, (0.86) | -0.02 (-0.20; 0.17) | 0.835 | NA |
| Pre-term birth (<37 weeks) ('no') | 737 (18.83%) | 997 (18.14%) | 689 (93.49%) | 931 (93.38%) | 1.02 (0.71; 1.47) | 0.894 | 0.00 (-0.02; 0.02) |
| Emergency Caesareans section ('no') | 765 (15.75%) | 1031 (15.35%) | 616 (80.52%) | 857 (83.12%) | 0.85 (0.57; 1.26) | 0.365 | -0.02 (-0.04; 0.03) |
| Low birth weight ('no') | 567 (37.56%) | 719 (40.97%) | 526 (92.77%) | 672 (93.46%) | 0.93 (0.54; 1.59) | 0.747 | -0.01 (-0.03; 0.03) |
| Smoking at discharge ('no') | 644 (29.85%) | 909 (26.10%) | 605 (93.94%) | 834 (91.75%) | 1.46 (0.61; 3.51) | 0.328 | 0.03 (-0.5; 0.11) |
| Initiated breastfeeding ('yes') | 680 (25.93%) | 931 (24.31%) | 613 (90.15%) | 829 (89.04%) | 1.22 (0.81; 1.84) | 0.295 | 0.01 (-0.02; 0.05) |

#### b) Main results for primary and secondary outcomes for follow-up 2 cohort

Results followed a similar pattern in the follow-up 2 cohort in which outcomes were measured three months after the intervention had ended (Table 2). There was a similar rate of initiation of antenatal care by 12 weeks and 6 days of pregnancy in both the intervention and control arm and there were no statistically significant differences by trial arm in any of the secondary outcomes.

#### c) Sub-group analysis

Pre-specified subgroup analyses of the primary outcome were conducted for parity, ethnicity, deprivation, cluster baseline rate of booking and implementation model. There was no evidence that the effect of the intervention differed by any of the variables considered (Table 3).

**Table 3:** Subgroup analysis of primary outcome.

|  | Intervention<br>N(% missing) | Control N (%<br>missing) | Odds ratio (95%<br>CI) | p-value for<br>interaction |
| --- | --- | --- | --- | --- |
| Parity of mother **) |  |  |  | 0.982 |
| 0 | 264 (0%) | 343 (0.29%) | 1.20 (0.64; 2.27) | n/a |
| 1+ | 588 (1.34%) | 686 (1.15%) | 1.14 (0.88; 1.47) | n/a |
| BL rate of booking |  |  |  |  |
| <70% | 405 (1.22%) | 551 (1.08%) | 1.12 (0.74; 1.68) | n/a |
| >=70% and <90% | 634 (0.94%) | 692 (0.57%) | 1.04 (0.78; 1.37) | n/a |
| Ethnicity ***) |  |  |  | 0.600 |
| White British | 169 (0%) | 169 (0%) | 1.24 (0.15; 9.93) | n/a |
| White (other) | 162 (1.38%) | 239 (1.01%) | 0.88 (0.52; 1.49) | n/a |
| Asian | 170 (0.58%) | 190 (1.04%) | 0.76 (0.27; 2.12) | n/a |
| Black | 101 (0%) | 183 (0%) | 0.93 (0.48; 1.79) | n/a |
| Other | 97 (3.00%) | 106 (0.93%) | 1.53 (0.77; 3.04) | n/a |
| Deprivation quintile *) |  |  |  | 0.310 |
| 1-2 | 77 (0%) | 241 (0.82%) | 1.17 (0.21; 6.48) | n/a |
| 3-4 | 323 (0.62%) | 432 (1.14%) | 1.03 (0.62; 1.72) | n/a |
| 5-6 | 364 (1.09%) | 322 (0.92%) | 0.83 (0.39; 1.76) | n/a |
| 7-8 | 114 (4.20%) | 189 (0%) | 0.99 (0.18; 5.35) | n/a |
| 9-10 | 78 (0%) | 43 (0%) | 1.85 (0.39; 8.73) | n/a |
| Intervention implementation |  |  |  | 0.595 |
| *Model A* (more focus on<br>quantity of conversations, less<br>focus on community<br>development) | 701 (0.99%) | 772 (1.03%) | 1.09 (0.84; 1.40) | n/a |
| *Model B* (less focus on<br>quantity of conversations,<br>more focus on community<br>development) | 338 (1.17%) | 471 (0.42%) | 1.00 (0.89; 1.28) | n/a |

\*) only 4 ward pairs used \*\*) only 6 ward pairs used \*\*\*) only 7 ward pairs use

#### d) Safety analysis

Safety analysis showed fewer maternal and infant deaths in the intervention clusters (see supplementary file 3).

## Discussion

This study succeeded in developing and implementing co-produced place-based interventions to increase early antenatal care initiation in 10 ethnically and linguistically diverse inner-city neighbourhoods. This was rigorously evaluated through a large-scale cluster randomised trial. The intervention did not show differences between the trial arms at either of the two follow-up time points in the primary outcome of accessing antenatal care before 12 completed weeks of pregnancy. The secondary outcomes also showed limited differences although the proportion of women who accessed care by 10 weeks was higher and antenatal admissions were lower in the intervention arm at the first follow-up whilst the intervention was running.

In considering why there was limited effect, our process evaluation pointed to several issues [24]. The interventions were dependent on the trickle down of messaging through community assets as well as direct contact with those in the neighbourhood. A focus on numbers of conversations by those commissioning and delivering the intervention meant that in some areas there was insufficient resource to put into fostering relationships with community assets, whilst in other areas the reverse applied. Running the intervention for longer than six months with bigger teams and including organisational development to support asset-based working may be needed to increase intervention strength for changing the socio-cultural environment to support early access to antenatal care. Similarly, our intervention may not have sufficiently reached those who were most at risk of late initiation of care, despite using local information in each site to identify key target groups.

The current staffing crisis in midwifery, combined with austerity of healthcare budgets in the UK, has placed additional limits within maternity services. Our intervention may have created extra demand for earlier initiation of antenatal care, which services may not have been able to meet. Recent systematic reviews on engagement with antenatal care have pointed to the importance of addressing structural and organisational barriers for deeply entrenched health inequities.

Recommendations from Sharma et al [14] for structural adaptations for accessible antenatal services for ethnically minoritised groups included the geographical location of services, the availability of interpreters and anti-racist and culturally inclusive practice training. These types of changes were beyond the scope of our intervention.

A large-scale randomised controlled trial requires rigour which was at times difficult to marry with the flexibility required in a co-produced community engagement intervention. Similar constraints have been expressed by others [29-31]. We found that our unit of randomisation – electoral wards – set up artificial geographical boundaries, which did not always reflect the neighbourhood areas defined by the communities with which we engaged. Indeed, their social networks often transcended geographical boundaries. From a community development perspective wards are relatively large geographical areas; in smaller ‘hyper-local’ neighbourhoods, community networks to stimulate change can be more easily developed.

Despite the limited positive results for this specific intervention, this trial does offer learning that those developing and implementing interventions to address health inequalities in health and care services can take forward regarding co-production of place-based community interventions [32]. In carrying out this work to strengthen community support for early care initiation we brought together and helped forge relationships between the UK National Health Service (NHS) and the wider community, across 10 areas at a time when working was very siloed. We found, however, that the absence of structures in place for engagement meant these processes often had no prior foundation and required more time and resource to develop impactful relationships. Such findings tally with those of South and colleagues in their study of the community champion model which rose in prominence at the height of the COVID-19 pandemic for its ability to connect with groups who were disproportionately impacted [33,34]. They also noted that a supportive infrastructure, training in community engagement and development of long-term community relationships were required.

This is a time of system transformation in the UK NHS with new Integrated Care Systems providing greater recognition and incentives for services to work with the community to reduce health inequalities [35]. This, and similar policy drivers in other countries towards community asset-based ways of working, could create better opportunities for embedding place-based interventions to strengthen community support for addressing health inequalities in maternity care in the future.

## Data Availability

All data produced in the present study are available upon reasonable request to the authors

https://osf.io/cgfjw/

## Acknowledgements

The authors would like to thank the staff in the NHS trust informatics teams who provided routine maternity data and all the community organisations, practitioners and members of the public who have assisted with the study. They would also like to thank: Inderjeet Kaur, Khalid Kahn, Joanne Oliber, Sandra Reading, Cathy Falvey-Brown, Sarah Latham, Kanta Patel, Belinda Green for their role in the early development of this research, obtaining funding and/or for facilitating the research at their NHS Trusts; staff at the former social design agency Uscreates and members of the Well Communities community engagement team at the University of East London for their work on the co-design of the intervention; and members of the data management team at Queen Mary University of London PCTU who advised on all data management matters. Angela Harden is supported by the NIHR North Thames Applied Research Collaboration (NT-ARC) held by Bart’s Health NHS Trust. The views expressed are those of the author(s) and not necessarily those of the NHS, the NIHR or the Department of Health and Social Care.

## Footnotes

1 Electoral wards are ‘used to elect local government councillors in metropolitan and non-metropolitan districts, unitary authorities and the London boroughs in England’. While population size varies, the English average is about 8,200 people per ward [36]

## References

1. Knight, M, Bunch, K, Tuffnell, D, Jayakody, H, Shakespeare, J, Kotnis, R, Kenyon, S & Kurinczuk, J (eds) 2018, Saving Lives, Improving Mothers’ Care - Lessons learned to inform maternity care from the UK and Ireland Confidential Enquiries into Maternal Deaths and Morbidity 2014-16. National Perinatal Epidemiology Unit: University of Oxford, Oxford. <https://www.npeu.ox.ac.uk/assets/downloads/mbrrace-uk/reports/MBRRACE-UK%20Maternal%20Report%202018%20-%20Web%20Version.pdf>

2. World Health Organization. WHO recommendations on antenatal care for a positive pregnancy experience. Geneva, Switzerland, 2016.

3. Partridge S, Balayla J, Holcroft CA, Abenheim HA. Inadequate prenatal care utilization and risks of infant mortality and poor birth outcome: a retrospective analysis of 28,729,765 U.S. deliveries over 8 years. Am J Perinatol. 2012;29:787–94.

4. Raatikainen K, Heiskanen N, Heinonen S. Under-attending free antenatal care is associated with adverse pregnancy outcomes. BMC Public Health. 2007; 10.1186/1471-2458-7-268.

5. National Institute for Health and Clinical Excellence. Antenatal Care for Uncomplicated Pregnancies: NICE guidelines [CG62]. Manchester: National Institute for Health and Clinical Excellence; 2021.

6. Cresswell J, Yu G, Hatherall B, Morris J, Jamal F, Harden A, Renton A. Predictors of the timing of initiation of antenatal care in an ethnically diverse urban cohort in the UK. BMC Pregnancy Childbirth. 2013; 13:103.

7. Downe S, Finlayson K, Walsh D, Lavender T. Weighing up and balancing out’: a meta-synthesis of barriers to antenatal care for marginalised women in high-income countries. BJOG. 2009;116:518–29.

8. Hatherall B, Morris J, Jamal F, Sweeney L, Wiggins M, Kaur I, et al. Timing of the initiation of antenatal care: an exploratory qualitative study of women and service providers in East London. Midwifery. 2016;36:1–7.

9. Henderson J, Gao H, Redshaw M. Experiencing maternity care: the care received and perceptions of women from different ethnic groups. BMC Pregnancy Childbirth. 2013; 10.1186/1471-2393-13-196. Accessed 30 Jan 17.

10. Hollowell J, Oakley L, Vigurs C, Barnett-Page E, Kavanagh J, Oliver S. Increasing the early initiation of antenatal care by black and minority ethnic women in the United Kingdom: a systematic review and mixed methods synthesis of women’s views and the literature on intervention effectiveness. Oxford: National Perinatal Epidemiology Unit; 2012.

11. Lindquist A, Kurinczuk JJ, Redshaw M, Knight M. Experiences, utilisation and outcomes of maternity care in England among women from different socio-economic groups: findings from the 2010 National Maternity Survey. BJOG. 2015;122(12):1610–7.

12. Oakley L, Gray R, Kurinczuk JJ, Brocklehurst P, Hollowell J. A systematic review of the effectiveness of interventions to increase the early initiation of antenatal care in socially disadvantaged and vulnerable women. Oxford: National Perinatal Epidemiology Unit; 2009.

13. Dada S, Tunçalp Ö, Portela A, Barreix M, Gilmore B. (2021) Community mobilization to strengthen support for appropriate and timely use of antenatal and postnatal care: a review of reviews. Journal of Global Health 11.

14. Sharma E; Pei-Ching Tseng; Angela Harden; Leah Li; Shuby Puthussery 2023 Ethnic minority women’s experiences of accessing antenatal care in high income European countries: a systematic review BMC Health Services Research

15. Ganju S, Khanna R, Taparia M, Hardikar N. Promoting accountability for maternal health through report card. COPASAH Communique. 2014;8:3–5

16. Carlo WA, Goudar SS, Jehan I, Chomba E, Tshefu A, Garces A, et al. Newborn-care training and perinatal mortality in developing countries. N Engl J Med. 2010;362:614–23.

17. McGowan, V.J., Buckner, S., Mead, R. et al. Examining the effectiveness of place-based interventions to improve public health and reduce health inequalities: an umbrella review. BMC Public Health 21, 1888 (2021). 10.1186/s12889-021-11852-z

18. Popay J, Halliday E, Mead R, Townsend A, Akhter N, Bambra C, et al. Investigating health and social outcomes of the Big Local community empowerment initiative in England: a mixed method evaluation. Public Health Res 2023;11(9)

19. Whitehead M A typology of actions to tackle social inequalities in health Journal of Epidemiology & Community Health 2007;61:473–478.

20. Morales-Garzón S, Parker LA, Hernández-Aguado I, González-Moro Tolosana M, Pastor-Valero M, Chilet-Rosell E. Addressing Health Disparities through Community Participation: A Scoping Review of Co-Creation in Public Health. Healthcare (Basel). 2023;11(7):1034. doi: 10.3390/healthcare11071034. PMID: 37046961; PMCID: PMC10094395.

21. Brunton G, Caird J, Kneale D, Thomas J, Richardson M (2015) Review 2: Community engagement for health via coalitions, collaborations and partnerships: a systematic review and meta-analysis. London: EPPI-Centre, Social Science Research Unit, UCL Institute of Education, University College London.

22. O’Mara-Eves A, Brunton G, McDaid D, Oliver S, Kavanagh J, Jamal F, et al. Community engagement to reduce inequalities in health: a systematic review, meta-analysis and economic analysis. Public Health Res 2013;1(4)

23. Sawtell, M., Sweeney, L., Wiggins, M. et al. Evaluation of community-level interventions to increase early initiation of antenatal care in pregnancy: protocol for the Community REACH study, a cluster randomised controlled trial with integrated process and economic evaluations. Trials 19, 163 (2018).

24. Sweeney L, Salisbury C, Wiggins M et al. (forthcoming) Factors influencing the implementation of a co-designed and co-delivered place-based intervention to strengthen community support for early initiation of antenatal care: findings from the Community REACH process evaluation.

25. Kutsnetova E, Hunter R, Harden A et al (forthcoming) Economic evaluation of a co-designed and co-delivered place-based intervention to strengthen community support for early initiation of antenatal care: findings from the Community REACH trial.

26. Findlay, G. and Tobi, P. 2017. Well communities. Perspectives in Public Health. 137 (1), pp. 17–20. 10.1177/1757913916680329

27. Hoffmann TC, Glasziou PP, Boutron I, Milne R, Perera R, Moher D, Altman DG, Barbour V, Macdonald H, Johnston M, et al. Better reporting of interventions: template for intervention description and replication (TIDieR) checklist and guide. BMJ. 2014;348:g1687

28. Norton, E. C., Miller, M. M., & Kleinman, L. C. (2013). Computing Adjusted Risk Ratios and Risk Differences in Stata. The Stata Journal, 13(3), 492–509. 10.1177/1536867X1301300304

29. Goodkind J.R., Amer S, Christian C, Hess, JM, Bybee D, Isakson, BL, Baca B, Ndayisenga M, Greene RN, Shantzek C. (2017) Challenges and Innovations in a Community-Based Participatory Randomized Controlled Trial. Health Education and Behaviour, Vol 44(1): 123–130.

30. McConnell, T., Best, P., Davison, G., McEneaney, T., Cantrell, C., Tully, M. (2018) Coproduction for feasibility and pilot randomised controlled trials: learning outcomes for community partners, service users and the research team. BMC Research Involvement and Engagement; 4:32.

31. Moore, G. F., Audrey, S., Barker, M., Bond, L., Bonell, C., Hardeman, W., Moore, L., O’Cathain, A., Tinati, T., Wight, D., & Baird, J. (2015). Process evaluation of complex interventions: Medical Research Council guidance. BMJ (Clinical research ed.), 350, h1258.

32. Salisbury, C, Harden A, Sweeney L et al (forthcoming) Development of a fidelity framework for co-producing interventions: Learning from the Community REACH trial.

33. South J, Bagnall AM, Jones R, Passey A, Woodall J, Gledhill R, Mapplethorpe T, Stansfield J (2021) Community champions: A rapid scoping review of community champion approaches for the pandemic response and recovery. London: Public Health England.

34. South, J., Woodall, J., Stansfield, J. et al. A qualitative synthesis of practice-based learning from case studies on COVID community champion programmes in England, UK. BMC Public Health 24, 7 (2024). 10.1186/s12889-023-17470-1

35. NHS England. 2023. What are integrated care systems? https://www.england.nhs.uk/integratedcare/what-is-integrated-care/

36. Office for National Statistics. 2024. Electoral wards and Divisions. (online) https://www.ons.gov.uk/methodology/geography/ukgeographies/administrativegeography/england#:~:text=Electoral%20ward%20and%20electoral%20division,divisions%2C%20with%206%2C904%20in%20England

