## Supplemental Table 1 for "Co-designed and co-delivered place-based community interventions to reduce inequity in early initiation of antenatal care: Findings from the cluster randomised controlled Community REACH trial"

**Table S1: Description of the Community REACH intervention using the TIDieR reporting template**

| **1. Brief Name** | **Community REACH** |
| --- | --- |
| **2. Why** | Rates of initiation of antenatal care by 12+6 weeks of pregnancy below national targets especially amongst under-served groups. Community REACH aims to a) raise awareness in local communities of the value of antenatal care and its early uptake and b) mobilise community and system support for women in how and when to access care and to change local social norms to sustain that support. It focuses on communities within local urban areas which report the highest rates of late initiation of care. |
| **3. What (procedures)** | The key steps followed in the co-design, set up and delivery of the intervention in each intervention site were:   1. ***Development of community profiles and asset mapping*** including stakeholder engagement and resident conversations through street engagement 2. ***Co-design workshops*** to inform intervention, communication strategy and materials 3. ***Recruitment of community organisation cohost*** including project co-ordinator 4. ***Intervention set-up***: a) recruitment and training of local community REACH volunteers and b) community cohost and REACH volunteers in each site (the community team) create local project plan including outreach plan, further development of local profiles and asset mapping, setting up local advisory group, holding launch event 5. ***Intervention delivery***: Community team carry out intervention delivery, including completion of conversation forms and monthly reports 6. ***Initial training and ongoing support*** for community teams including reflective working together workshops |
| **4. What (materials)** | 1. ***Development of community profiles and asset mapping:*** phone and computer access for desk top research and stakeholder engagement; maps and conversation record forms for street engagement 2. ***Co-design workshops:*** ideas boards, proto-personas 3. ***Recruitment of community organisation cohost:***  application form and criteria 4. ***Intervention set-up***: co-host and volunteer intervention toolkit and information pack; launch event venue and materials 5. ***Intervention delivery***: co-designed information leaflets and conversation cards, promotional materials (T-shirts, badges, pens, tote bags) 6. ***Initial training and ongoing support:*** facilitation activities, presentations, feedback reports, certificates for REACH volunteers |
| **5. Who** | 1. ***Development of community profiles and asset mapping:***  Research team; community engagement team 2. ***Co-design workshops:*** Community engagement team 3. ***Recruitment of community organisation cohost:*** Community engagement lead and project manager 4. ***Intervention set-up***: Co-host project manager and project co-ordinator; volunteers 5. ***Intervention delivery***: Co-host project manager and project co-ordinator; volunteers 6. ***Initial training and ongoing support:*** Community engagement team |
| **6. How** | Community REACH co-ordinator and volunteer team engaged in person with their local communities about antenatal care, through outreach activities, group presentations and one to one sessions |
| **7. Where** | The intervention was conducted in 20 ethnically and linguistically diverse inner-city electoral wards.  Intervention activities took place within ward boundaries in local community settings, at community events/activities, evening classes, faith groups; in places of high footfall e.g. GP surgeries, pharmacies, shopping centres; and informal, opportunistic outreach on the street |
| **8. When and how much** | The intervention was delivered over a six-month period with each Community REACH volunteer typically spending at least 1-2 hours per week doing outreach work. Each co-host aimed to recruit at least 10 volunteers. Community REACH project co-ordinators were contracted to undertake intervention activities 2 days per week. |
| **9. Tailoring** | Through the co-production process the components of the intervention were tailored to the local community to address cultural beliefs and motivational barriers and strengthen the appropriateness of the intervention for each intervention site e.g. volunteer team, outreach activities |
| **10. Modifications** | Delivering intervention messages in languages other than English.  Less focus on conversation targets.  Pairing up volunteers.  Letter to key stakeholders to raise awareness of intervention.  Flexibility for local advisory group format and timing of ‘launch event’ for intervention in each area.  End of project thank you event and volunteer certificate. |
| **11. How well: planned** | The Community REACH trial theory of change model was used to guide assessment of intervention fidelity and a set of 14 ‘operational deliverables’ were drawn up covering intervention set up (e.g. draw up a local outreach plan, recruit at least 10 volunteers); intervention delivery (e.g. convene and hold three local advisory group meetings; support each volunteer to carry out at least 50 conversations per month) and co-production (e.g. Work collaboratively with volunteers to deliver the intervention and to develop new skills and competences to advance their personal goals; work with local stakeholders to ensure intervention messages are embedded at all levels within the local community) . Fidelity data was collected through monthly meetings, monthly monitoring reports, observations and qualitative interviews. An assessment was also made of the extent to which the intervention in each site focused on wider community development work as well as conversation with residents. |
| **12. How well: actual** | Six out of the ten intervention sites showed medium to high fidelity to the operational deliverables whilst fours sites showed low fidelity. |
