## Supplemental Table 2 for "Co-designed and co-delivered place-based community interventions to reduce inequity in early initiation of antenatal care: Findings from the cluster randomised controlled Community REACH trial"

**Table S2: Sensitivity analysis of primary outcome**

|  | **Included in analysis** | | **Treatment effect** | | |
| --- | --- | --- | --- | --- | --- |
|  | **Intervention N (%)** | **Control N (%)** | **OR (95% CI)** | **OR p-value** | **RD (95% CI)** |
| Per protocol analysis | 924 (88.0%) | 1088 (86.8%) | 1.05 (0.85; 1.29) | 0.604 | 0.01 (-0.01; 0.04) |
| Adjusted for ethnicity and IMD | 865 (82.4%) | 1102 (87.9%) | 1.08 (0.68; 1.72) | 0.711 | 0.01 (-0.06; 0.08) |
| **Imputation of missing primary outcomes** | | | | | |
| Substitution of all missing values of the outcome as having booked after 12 weeks + 6 days for both arms. | 1050 (100%) | 1253 (100%) | 1.06 (0.89; 1.26) | 0.480 | 0.01 (-0.01; 0.04) |
| Substitution of all missing values of the outcome as having booked after 12 weeks + 6 days in the intervention arm and before 12 weeks + 6 days in control arm wards*. | 1050 (100%) | 1253 (100%) | 0.99 (0.83; 1.20) | 0.950 | 0.01 (-0.02; 0.03) |
| Substitution of all missing values of the outcome as having booked before or after 12 weeks + 6 days with probability 0.5 in both arms | 1050 (100%) | 1253 (100%) | 1.06 (0.88; 1.28) | 0.478 | 0.01 (-0.01; 0.04) |

*This is the most extreme imputation option and provides a lower limit for potential effect estimates.
