## Supplemental Table 3 for "Co-designed and co-delivered place-based community interventions to reduce inequity in early initiation of antenatal care: Findings from the cluster randomised controlled Community REACH trial"

**Table S3: Safety analysis**

|  | **Baseline** | | **Follow-up 1** | | **Follow-up 2** | |
| --- | --- | --- | --- | --- | --- | --- |
|  | **Intervention (n=882)** | **Control (n=1089)** | **Intervention (n=969)** | **Control (n=1148)** | **Intervention (n=840)** | **Control (n=1143)** |
| Maternal death N(%) | 0 (0.00%) | 1 (0.09%) | 0 (0.00%) | 0 (0.00%) | 0 (0.00%) | 0 (0.00%) |
| Infant death N(%) | 3 (0.34%) | 3 (0.28%) | 3 (0.29%) | 9 (0.72%) | 2 (0.24%) | 3 (0.26%) |
